# Safety of Flexible Sigmoidoscopy in Pregnant Patients with known or suspected Inflammatory Bowel Disease

**DOI:** 10.1101/19007997

**Authors:** Myung S. Ko, Vivek A. Rudrapatna, Patrick Avila, Uma Mahadevan

## Abstract

**Background and Aims:** Lower gastrointestinal endoscopy is the gold standard for the diagnosis and staging of Inflammatory Bowel Disease (IBD). However, there is limited safety data in pregnant populations, resulting in conservative society guidelines and practice patterns favoring diagnostic delay. The aim of this study is to investigate if the performance of flexible sigmoidoscopy is associated with adverse events in pregnant patients with known or suspected IBD.

**Methods:** A retrospective cohort study was conducted at the University of California San Francisco (UCSF) between April 2008 and April 2019. Female patients aged between 18 and 48 years who were pregnant at the time of endoscopy were identified. All patient records were reviewed to determine disease, pregnancy course, and lifestyle factors. Two independent reviewers performed the data abstraction. Adverse events were assessed for temporal relation (defined as within 4 weeks) with endoscopy. Any discrepancies in the two reviewers’ data were reviewed by a third independent investigator. Descriptive statistics of data were calculated, and comparison of continuous and categorical data were made using a one-sided Wilcoxon rank-sum test and Fisher’s exact test, respectively.

**Results:** We report the outcomes of 48 pregnant patients across all trimesters who underwent lower endoscopy for suspected or established IBD. There were no hospitalizations or adverse obstetric events temporally associated with sigmoidoscopy. 78% of patients experienced a change in treatment following sigmoidoscopy. 12% of the patients with known IBD were found to have no endoscopic evidence of disease activity despite symptoms.

**Conclusions:** Lower endoscopy in the pregnant patient with known or suspected IBD is low risk and affects therapeutic decision making. It should not be delayed in patients with appropriate indications.

## Introduction

The pregnant woman with active inflammatory bowel disease (IBD) presents management challenges to the treating physician. Among women with IBD at the time of conception, 36% of Ulcerative Colitis (UC)^1^ and 33% of Crohn’s Disease (CD)^2^ patients will experience an increase in disease activity. Active disease is associated with increased risk of adverse pregnancy outcomes, including spontaneous abortion, preterm birth, and low birth weight.^3–4^ Accurately establishing the presence and severity of IBD activity during pregnancy in the symptomatic patient is a critical juncture in decision making.

There are several challenges in accurately establishing the presence and severity of IBD activity during pregnancy. Due to physiologic changes and alterations in gastrointestinal motility, pregnancy often presents with a myriad of gastrointestinal symptoms such as abdominal pain, constipation, hemorrhoids and diarrhea, that mimic symptoms of an IBD flare.^5^ The noninvasive modalities of assessing for evidence of inflammation – such as erythrocyte sedimentation rate, C-reactive protein – lose their predictive value in pregnancy.^6–7^ While levels of fecal calprotectin (FCP) have been shown to be elevated in pregnant patients with IBD, there is currently no established cut-off for FCP that effectively predicts disease activity in the pregnant IBD patients.^7^ Furthermore, several meta-analyses have suggested that there is a significant false positive rate, which in the pregnant patient would lead to inappropriate exposure to escalated IBD therapy.^8–10^ As such, an endoscopic evaluation is critical in accurately staging the severity of IBD in pregnancy and avoiding delay in care.

Potential risks associated with endoscopy during pregnancy include fetal hypoxia from increased intra-abdominal pressure during endoscopy. Hence, the American Society of Gastrointestinal Endoscopy (ASGE) currently recommends that endoscopy be postponed to second trimester whenever possible.^11^ However, delayed diagnostic work-up of a suspected IBD flare in a pregnant patient has serious consequences, as does empiric therapy with biologics and immunosuppressants.

Based on the paucity of studies studying the use of lower endoscopy in pregnancy, larger numbers are needed to fully understand the risks versus benefits of lower endoscopy in pregnant IBD patients. The aim of this study is to investigate if the performance of flexible sigmoidoscopy is associated with adverse events in pregnant patients with IBD.

## Methods

A retrospective cohort study was conducted at the University of California San Francisco (UCSF) between April 2008 and April 2019. Institutional ethics approval was obtained (IRB 18-26237). Structured EHR data was extracted from the UCSF Epic system using Clarity and Caboodle tools^7^. Prior to being used for this study, the data was de-identified to comply with the US Department of Health and Human Services ‘Safe Harbor’ guidance. This database was queried to identify female patients aged between 18 and 48 years inclusive bearing diagnosis codes corresponding to IBD, melena, hematochezia, or diarrhea and procedure billing codes corresponding to lower gastrointestinal endoscopy. Additional patients were separately identified from a database of endoscopic reports using keyword-based search (see Supplemental Methods). All records meeting the above criteria were subsequently assessed by two independent reviewers (MK, PA) to confirm pregnant status at the time of lower gastrointestinal endoscopy.

Eligible patient records were further reviewed to determine disease, pregnancy course, lifestyle factors such as smoking and alcohol use, bowel preparation, and pregnancy outcomes. All adverse events were assessed for temporal relation (defined as within 4 weeks) with endoscopy. Any discrepancies in the two reviewers’ data were further reviewed by a third independent investigator (VAR).

Descriptive statistics of continuous data were calculated as means and medians with standard deviation and interquartile ranges (IQR), respectively. Categorical data were reported by absolute numbers and percentages. Comparisons of continuous data were made using t-tests with unequal variance for normally-distributed data, or the Wilcoxon rank-sum test otherwise. Comparisons of categorical data were calculated using Fisher’s exact test.

## Results

In total, 48 pregnant patients underwent 47 flexible sigmoidoscopies and 3 colonoscopies. 41 (85%) of these patients had a previous diagnosis of IBD (7 Crohn’s Disease, 34 Ulcerative Colitis). Median maternal age was 33 years [IQR: 30.1-35] (Table 1). Sedation was used in 5/50 (10%) procedures and 86% had biopsies. Monitored anesthesia care was present in 3 cases (2 colonoscopy and 1 sigmoidoscopy). Median sigmoidoscopy scope insertion was 30 cm [range: 15-40]. A total of 8 endoscopies were performed in the first trimester, 26 in the second, and 16 in the third trimester.

**Table 1.** Baseline Characteristics.

|  |  |  |
| --- | --- | --- |
| **Type of Lower Endoscopy** | **Colonoscopy** | 3 (6.0%) |
|  | **Sigmoidoscopy** | 47 (94.0%) |
| **Diagnosis** | **Crohn's Disease** | 7 (14.5%) |
|  | **Ulcerative Colitis** | 34 (70.8%) |
|  | **Non-IBD** | 7 (14.6%) |
| **Median Age (years)** |  | 33<br>[30.1-35] |
| **Disease Location (Crohn's Disease)** | **Ileal** | 0 |
|  | **Colonic** | 3 (42.8%) |
|  | **Ileocolonic** | 4 (57.1%) |
|  | **Perianal fistula** | 1 (14.2%) |
| **Disease Location (Ulcerative Colitis)** | **Proctitis** | 4 (11.8%) |
|  | **Left-sided** | 19 (55.9%) |
|  | **Pancolitis** | 11 (32.4%) |
| **Median Gestational Age (weeks)** |  | 23<br>[1-36] |
| **Number of Procedures by Trimester** | **First Trimester** | 8 (16.0%) |
|  | **Second Trimester** | 26 (52.0%) |
|  | **Third Trimester** | 16 (32.0%) |
| **Sedation Use** |  | 5 (10.0%) |
| **Biopsies Obtained** |  | 43 (86.0%) |
| **Median Scope Insertion (cm)** |  | 30<br>[15-40] |

<sup>1</sup> Data presented as percent or median (IQR)

There were no hospitalizations or pregnancy adverse events temporally associated with flexible sigmoidoscopy in either IBD or non-IBD patient groups. One intrauterine fetal demise between 30-34 weeks gestational age (GA) (procedure at 10 weeks GA) and one elective termination in the second trimester (procedure at 13 weeks GA) occurred in the IBD group, but were not temporally or etiologically thought to be related to the sigmoidoscopy. Median GA at birth was 39 weeks in the IBD group [IQR 2] and 40.2 weeks in the non-IBD group [IQR 0.9, p=0.03]. Rates of caesarean section delivery were 32.4% and 16.7% in IBD and non-IBD women, respectively (Table 2, p=0.6).

**Table 2.**
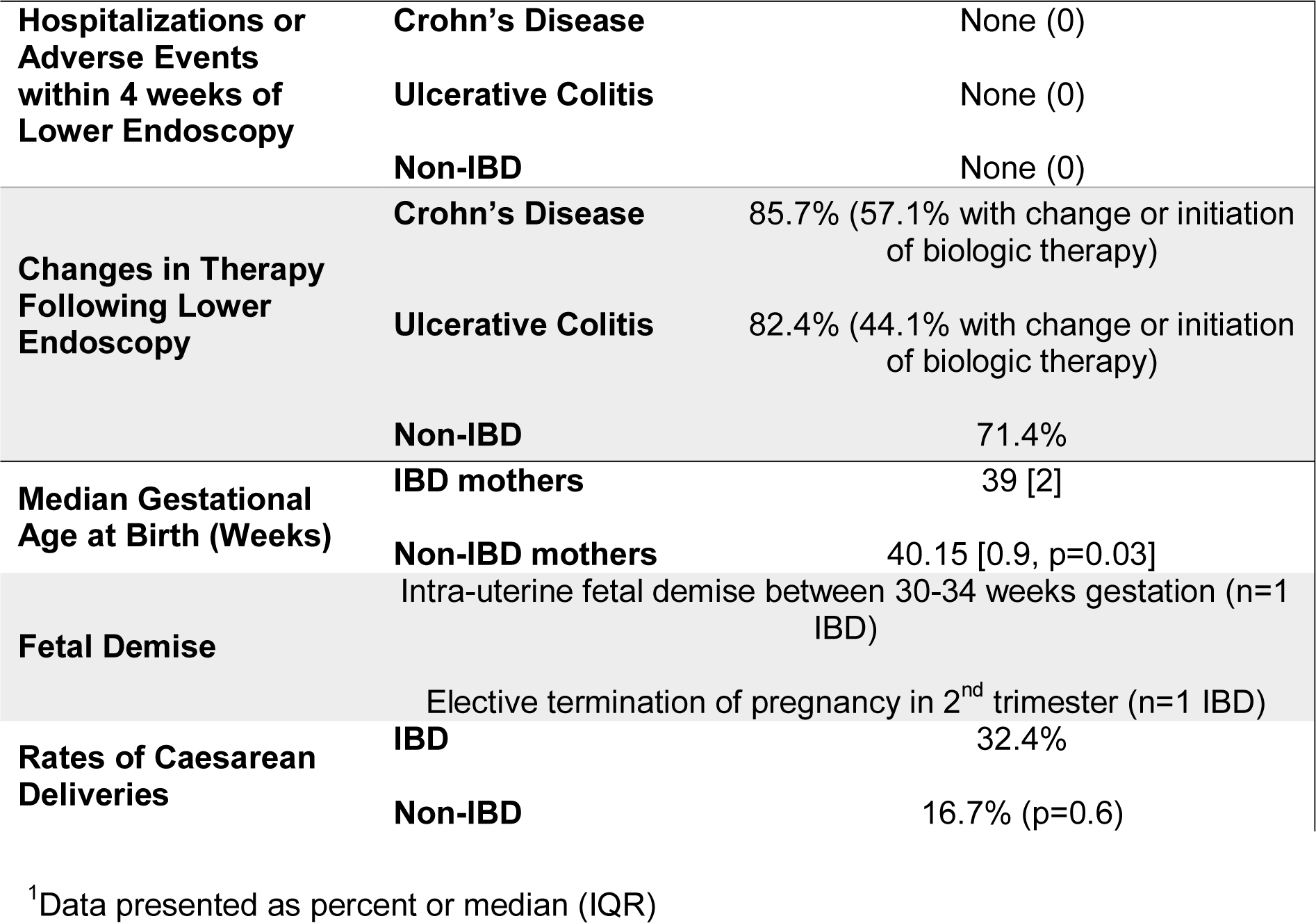
Adverse Events and Outcomes.

|  |  |  |
| --- | --- | --- |
| **Hospitalizations or Adverse Events within 4 weeks of Lower Endoscopy** | **Crohn's Disease** | None (0) |
|  | **Ulcerative Colitis** | None (0) |
|  | **Non-IBD** | None (0) |
| **Changes in Therapy Following Lower Endoscopy** | **Crohn's Disease** | 85.7% (57.1% with change or initiation of biologic therapy) |
|  | **Ulcerative Colitis** | 82.4% (44.1% with change or initiation of biologic therapy) |
|  | **Non-IBD** | 71.4% |
| **Median Gestational Age at Birth (Weeks)** | **IBD mothers** | 39 [2] |
|  | **Non-IBD mothers** | 40.15 [0.9, p=0.03] |
| **Fetal Demise** | Intra-uterine fetal demise between 30-34 weeks gestation (n=1 IBD) |  |
|  | Elective termination of pregnancy in 2<sup>nd</sup> trimester (n=1 IBD) |  |
| **Rates of Caesarean Deliveries** | **IBD** | 32.4% |
|  | **Non-IBD** | 16.7% (p=0.6) |

<sup>1</sup>Data presented as percent or median (IQR)

In the IBD group, 88.4% of women had evidence of disease activity at the time of flexible sigmoidoscopy, with 62.7% of women showing evidence of moderate to severe disease. For the 20 women who were found to have severe disease, 11 women (55%) were started on biologics for the first time, 1 woman (5%) underwent a switch in her biologic therapy, 6 women (30%) were started on systemic steroids, and 1 woman underwent a proctocolectomy during pregnancy (Table 3). Conversely, for the non-IBD pregnant patients who underwent a lower endoscopy, 5 patients (71.4%) were diagnosed with internal hemorrhoids, and the rest had normal endoscopic findings. No incident cases of IBD were diagnosed in this group.

**Table 3.** Therapeutic Impact of Endoscopy Findings.

|  | **N (%)** | **Added**<br>**Systemic**<br>**Steroids** | **Increased**<br>**biologic dose** | **Started new**<br>**biologic therapy** | **Switched**<br>**biologic**<br>**Therapy** |
| --- | --- | --- | --- | --- | --- |
| **Total** | 43 | 7 (16.3) | 1 (2.3) | 16 (37.2) | 1 (2.3) |
| **Remission** | 5 (11.6) | 0 | 0 | 0 | 0 |
| **Mild** | 11(25.6) |  |  | 3 (27.3) | 0 |
| **Moderate** | 7 (16.2) | 1(14.3) | 0 | 2 (28.6) | 0 |
| **Severe** | 20 (46.5) | 6 (30) | 1 (5) | 11 (55) | 1 (5) |

<sup>1</sup> Data presented as absolute number and percent

## Discussion

The mainstay of caring for women with IBD of childbearing age is careful preconception counseling and optimization of medical therapy to ensure steroid free clinical remission of at least 3 months prior to conception.^3^ Despite best efforts, however, many women with IBD experience worsening of their disease activity during pregnancy. The current ASGE guidelines recommend deferring lower endoscopy to second trimester or to the post-partum period; however, risks of IBD disease activity to the pregnancy call for timely endoscopic evaluation. This is why the American Gastroenterology Association Pregnancy Care Pathway recommends unsedated, unprepped flexible sigmoidoscopy as needed in any trimester in the patient with IBD.^3^

The safety of endoscopy was previously studied in a nationwide population-based cohort study by Ludvigsson et al.^12^ This studied 3052 pregnancies exposed to upper endoscopies, lower endoscopies, or endoscopic retrograde cholangiopancreatographies, and showed that exposure to any endoscopic procedure during pregnancy was associated with an increased risk of preterm birth and small for gestational age, independent of trimester. However, studies specifically studying the pregnant IBD patient population, which has increased rates of spontaneous abortions and pregnancy-related complications than their non-IBD counterparts, have been few and small in size. A matched case-control study of 42 IBD patients published by De Lima et al.^13^ reported two spontaneous abortions that were thought to be temporally related to lower gastrointestinal endoscopy. However, the authors noted that the spontaneous abortion rate was higher in the control population of IBD patients not undergoing endoscopy, and overall reported no increased adverse outcomes for the mother or fetus in any trimester. Consistent with prior studies, our work – the largest study to date of pregnant patients with known or suspected IBD undergoing flexible sigmoidoscopy – supports the safety of flexible sigmoidoscopy as a diagnostic procedure for patients with strong indications. An unsedated, unprepped flexible sigmoidoscopy without obstetric monitoring was found to be low risk and not associated with any maternal or fetal adverse events. These results reflect the combined experience of 11 different endoscopists and capture all trimesters of pregnancy. In total, approximately 78% of patients experienced a change in their therapy following the lower endoscopy.

In higher-risk groups such as pregnant patients presenting with red-flag symptoms, the decision to proceed with diagnostic lower endoscopy is frequently contrasted with non-invasive alternatives such as fecal calprotectin. Although the fecal calprotectin is a commonly-used biomarker for IBD, its sensitivity and specificity are insufficiently precise to be used as a primary diagnostic tool for IBD^7–10,14^, let alone to make major therapy changes in the gravid patient. We note that 15% of the patients in this cohort had “negative” lower endoscopic findings. Changes in bowel habits and hemorrhoids are common in the pregnant patient, and these results highlight the importance of confirming the diagnosis prior to initiation or escalation of immunosuppression.

In line with previously published data, pregnant IBD women in this cohort had higher rates of Cesarean section delivery than their non-IBD counterparts. This finding is similar to the results of large population-based studies that showed that women with IBD have a 1.5 to 2-fold increase in the rate of Cesarean delivery.^15–17^ The pregnant IBD women were also noted to have a slight reduction in median gestational age at birth by 1 week compared to their non-IBD counterparts. While this was a statistically significant difference, it is not a clinically significant one - both groups of women had a median gestational age of at least 39 weeks, a clinical timepoint after which adverse neonatal outcomes are seen to decrease.^17^

This study is limited by its sample size and retrospective nature. The absolute risk of adverse events following sigmoidoscopy in the general population is small, and as such our study is insufficiently powered to estimate these risks in this special population. We cannot exclude the possibility of confounding bias relevant to the decision of which patients should undergo lower endoscopy. Moreover, we note that this study was carried out at a tertiary care medical center with expertise in both IBD and Obstetrics.

In spite of these limitations, this study contributes significant additional case experience to the literature on this understudied and vulnerable population. Although pregnancy is a common phenomenon in the general population, interventional clinical studies in this group are rare precisely because of the risks – whether perceived or real. Although this attitude reflects an appropriate sensitivity towards this group, the gaps in knowledge and risk-adverse climate in clinical practice can paradoxically lead to overly conservative guidance and lower-quality care. In the absence of controlled trials, we suggest that cohort studies such as this are essential to demystify the perceived risks of medically necessary procedures and increase awareness and engagement by both the clinical and research communities. We hope subsequent guidelines will reflect this approach.

In conclusion, this study adds to the growing body of data supporting the safety of lower endoscopy in pregnant patients presenting with signs and symptoms concerning for new-onset or worsening Inflammatory Bowel Disease. Lower endoscopy in women with IBD during all trimesters of pregnancy is of low risk for mother and child. When indicated, this procedure should not be deferred as it directly impacts the medical decision making needed for optimal obstetric and gastrointestinal outcomes.

## Data Availability

There are no plans to make the data available relevant to this manuscript.

## Acknowledgements

The authors thank the UCSF Academic Research Systems unit including Dana Ludwig and Boris Oskotsky for electronic health record database generation and management. They also thank members of the UCSF Division of Gastroenterology for valuable discussion and feedback on this work.

## Conflicts of Interest

None from all authors

## Financial Support

VAR was supported by the National Institute of Diabetes and Digestive and Kidney Disease of the National Institutes of Health under award number T32 DK007007-42. UM was supported by a senior research award from the Crohn’s Colitis Foundation of America.

## Supplemental Methods

### EHR Structured Database Search Criteria

1. 17 < Age < 49, Female Gender
2. Having at least one of the following ICD10 (K50*/K51*/K52*/R19.7/K59.1/K58.0/K62.5) or ICD9 (555*/556*/558*/569.3/578.1/787.91/564.5/564.1) codes ever in their chart
3. Having any of the following CPTs for sigmoidoscopy, proctosigmoidoscopy, pouchoscopy, ileoscopy, or colonoscopy: ‘45330’, ‘45331’, ‘45332’, ‘45333’, ‘45334’, ‘45335’, ‘45336’, ‘45337’, ‘45338’, ‘45339’, ‘45340’, ‘45341’, ‘45342’, ‘45343’, ‘45344’, ‘45345’, ‘45346’, ‘45347’, ‘45348’, ‘45349’, ‘45350’, ‘G0104’, ‘G0106’, ‘45300’, ‘45303’, ‘45305’, ‘45307’, ‘45308’, ‘45309’, ‘45315’, ‘45315’, ‘45317’, ‘45320’, ‘45321’, ‘45327’, ‘44385’, ‘44386’, ‘44380’, ‘44381’, ‘44382’, ‘44383’, ‘44384’, ‘45378’, ‘45379’, ‘45380’, ‘45381’, ‘45382’, ‘45383’, ‘45384’, ‘45385’, ‘45386’, ‘45387’, ‘45388’, ‘45389’, ‘45390’, ‘45391’, ‘45392’, ‘45393’, ‘45394’, ‘45395’, ‘45396’, ‘45397’, ‘45398’, ‘G0121’, ‘G0121-53’, ‘G0122’

### Endoscopy Report Database Search Criteria

Keyword search terms: “pregnancy”, “pregnant”, “conception”, “gestation”, “obstetric”

## Notes

### Competing Interest Statement

The authors have declared no competing interest.

### Author Declarations

All relevant ethical guidelines have been followed and any necessary IRB and/or ethics committee approvals have been obtained.

Any clinical trials involved have been registered with an ICMJE-approved registry such as ClinicalTrials.gov and the trial ID is included in the manuscript.

